# Persistent Anthropometric Deficits in School-aged Children with Perinatal HIV Exposure

**DOI:** 10.64898/2026.04.07.26349779

**Authors:** Fatimah Donaldson, David Morgenthal, Amy Davidow, Jibreel Jumare, Paul Akhigbe, Esosa Osagie, Augustine Omoigberale, Ozoemene Obuekwe, Praise Okoh-Aihe, DOMHaIN study team, Vincent Richards, Modupe Coker

## Abstract

**Background:** Despite scale-up of antiretroviral therapy (ART), children living with HIV (CLHIV) and children who are HIV-exposed-but-uninfected (CHEU) are at an increased risk of poor growth outcomes compared to children HIV-unexposed-and-uninfected (CHUU). Few studies quantify the magnitude of growth deficits extending into school age in sub-Saharan Africa (SSA). This study examined the impact of perinatal HIV exposure and infection on the growth trajectory of school-aged children in Nigeria.

**Methods:** Within a prospective cohort, 569 children aged 3-11 years were recruited from pediatric clinics in Nigeria and matched by age and sex based on their exposure or infection status. School-aged children were observed across three time-points at 6-month intervals, during which anthropometric measures, CD4 count, and maternal factors were collected. Z-scores for height-for-age (HAZ), weight-for-age (WAZ), and body-mass-index-for-age (BAZ) were calculated using WHO standards. Longitudinal linear regression analyses using generalized estimating equations (GEE), adjusted for maternal and child covariates, were conducted to compare growth outcomes across groups.

**Results:** Growth Z-scores declined until approximately age 8, after which they gradually increased. Across all visits, CLHIV consistently and independently demonstrated lower Z-scores (WAZ (β = -1.04, p <0.001); HAZ (β = -0.67, p <0.001)), followed by CHEU with intermediate but significant impairments (WAZ (β = -0.35, p <0.01); HAZ (β = -0.38, p <0.01)) compared to CHUU.

**Conclusion:** Stunting remains unacceptably high in CLHIV and CHEU in SSA. The findings suggest a need for immediate paradigm shifts to address persistent growth deficits despite ART and beyond infancy.

## Introduction

HIV/AIDS remains a major global epidemic, affecting 39 million people in 2022, including 1.5 million children under 15 years of age. Nigeria has one of the largest HIV-affected populations in sub-Saharan Africa (SSA), with 1.9 million people living with HIV, representing 9% of the global HIV burden^1,2^. Despite expanded antiretroviral therapy (ART) coverage, recent studies show persistent growth deficits among children who were HIV-exposed but uninfected (CHEU) in SSA^3,4^. An estimated 2.6 million children in the region are living with HIV, representing 85% of the total global pediatric HIV population^5^.

Early studies^6,7^ show that infants born to mothers living with HIV experience reduced growth during infancy, including low birth weights and impaired intrauterine growth^3,4,8,9^. Growth aberration is commonly observed in children living with HIV (CLHIV), where infection and socioeconomic factors compromise growth^10,11^. Malnutrition and HIV interact synergistically to contribute to stunting, where malnutrition can accelerate the progression of AIDS, due to the weakening of the immune system^11–13^. Stunting is particularly prevalent among CLHIV and CHEU, while children who were HIV-uninfected and unexposed (CHUU) show the least impairment^3,14–16^. Prior studies^3,17–21^ show post-natal differences between CHEU and CHUU, with maternal ART initiation and prenatal exposure influencing infant growth outcomes.

This prospective cohort study examines the impact of perinatal HIV exposure and infection on growth among school-aged children in Nigeria. The setting is particularly important given that Nigeria is one of the five countries that together account for over half of all children globally who are perinatally HIV-exposed^22^. By including three study groups: CLHIV, CHEU, and CHUU, our objective is to examine changes in anthropometric Z-scores over time, investigating how HIV exposure and infection, with maternal factors, influence growth trajectories. We hypothesized that, even in the era of expanded ART access, CLHIV and CHEU would exhibit suboptimal growth, reflected by lower anthropometric-Z-scores compared to CHUU; further anticipated that CLHIV would have the lowest anthropometric-Z-scores among the three study groups. Understanding the impact of perinatal HIV exposure +/- infection allows us to better characterize the mechanisms driving persistent growth deficits despite ART access. This knowledge can inform the development of targeted interventions that address the unique metabolic, immunological, and nutritional challenges faced by CLHIV and CHEU.

## Methods

### Study Population

This prospective cohort included children living with HIV (CLHIV), children who were HIV-exposed but uninfected (CHEU), and children who were HIV-uninfected and unexposed (CHUU) children aged 3-11 years. Participants were recruited between May and December 2019 from pediatric and HIV care and treatment clinics at the University of Benin Teaching Hospital (UBTH) in Edo state, Nigeria. Recruitment of mother-child pairs was conducted using convenience sampling, and participants were matched by age and sex into the *Dental Caries and its association with Oral Microbiomes and HIV in young children — Nigeria (DOMHaIN)* study^23^. A cohort investigating the relationship between the oral microbiome and HIV status. Questionnaires were administered at enrollment, and again at 6 months and 12 months. Informed consent/assent was obtained per institutional guidelines (University of Maryland, Baltimore (HP-00084081), Rutgers State University of New Jersey (Pro2019002047), and University of Benin Teaching Hospital, Benin City (ADM/E22/A/VOL. VII/14713)).

CLHIV and CHEU were recruited from the pediatric HIV care and treatment clinic or through referrals from mothers attending the adult ART clinic at UBTH. Children in the CHUU group were recruited from the hospital when attending well-child or pediatric clinics at UBTH or by referral from the communities of CLHIV and CHEU children. Premature deliveries were defined by gestational age of < 37 weeks^19^, obtained from medical records. Anthropometric z-scores, height-for-age (HAZ), weight-for-age (WAZ), and body-mass-index-for-age (BAZ) were computed using the *zscorer* package in R^24^. Weight-for-height (WFH) was excluded from inferential analyses due to >65% missingness. Malnutrition was classified according to WHO standards: stunting (HAZ < -2), underweight (WAZ < -2), and wasting (BAZ < -2). BAZ is used to assess wasting as the primary indicator due to the WHO recommendation of children ≤ 5 years for WFH and BAZ for older children. WFH scores were not included due to >65% missingness (387 observations).

### Statistical Analysis

All analyses were performed in R (version 4.3.2). Continuous variables were compared using ANOVA or the Kruskal-Wallis test, while categorical variables were compared using Chi-square or Fisher’s exact tests (p < 0.05). HAZ, WAZ, and BAZ were plotted by age for each HIV group with a line of best fit to apply to the three study groups.

Longitudinal analyses were performed using generalized estimating equations (GEE). Models adjusted for the visit number, sex, breastfeeding status, and delivery method; diet and maternal employment were explored but excluded due to poor model fit (high QIC). Dietary frequency was collapsed into three categories (rarely, weekly, and daily) to address sparse data.

The GEE approach and QIC-based selection prioritized models that better captured repeated measures. Model fit was evaluated using the quasi-likelihood under the independence model criterion (QIC), with a lower QIC value indicating a better fit. Regression coefficient and corresponding 95% confidence intervals were reported. P-value <0.05 was statistically significant.

To evaluate potential bias due to loss-to-follow-up, baseline characteristics of children who completed all visits were compared with those lost before the third visit using t-tests (continuous variables) and chi-square tests (categorical variables).

## Results

At baseline, 569 children (CLHIV: n= 190; CHEU: n= 189; CHUU: n= 190) from the DOMHaIN cohort study were included based on available data. Participants without a confirmed infection status were omitted from the analysis. Children were followed across three visits at six-month intervals; variation in participant numbers was due to loss-to-follow-up.

### Children and Maternal Baseline Characteristics and Comparisons

**Table 1** describes children’s and material demographics at baseline, categorized by HIV exposure or infection status. At enrollment, the ages of children ranged from 41 to 127 months. Mean ages of 7.28 years for CLHIV, 7.06 for CHEU, and 7.17 years for CHUU.

**Table 1.**
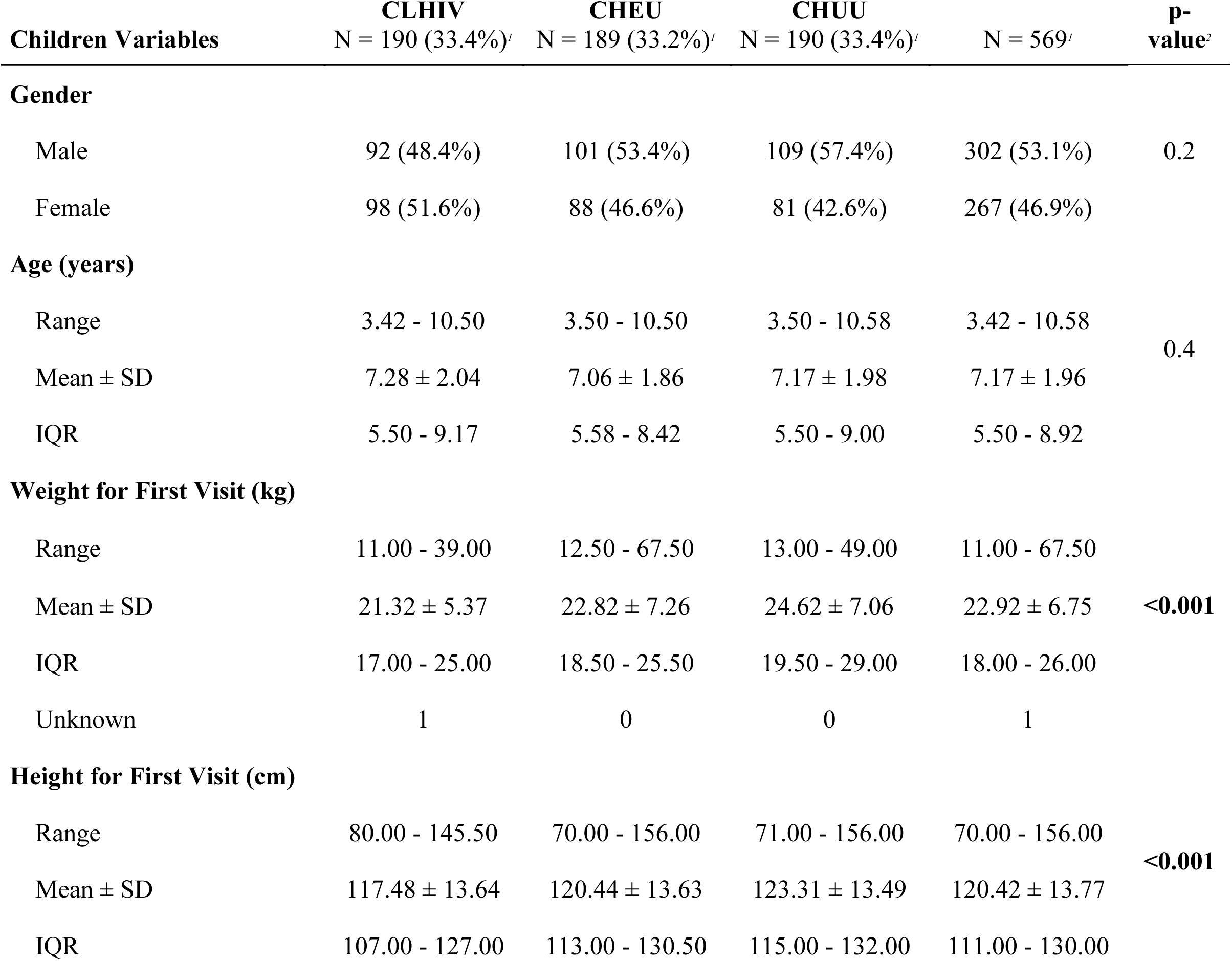
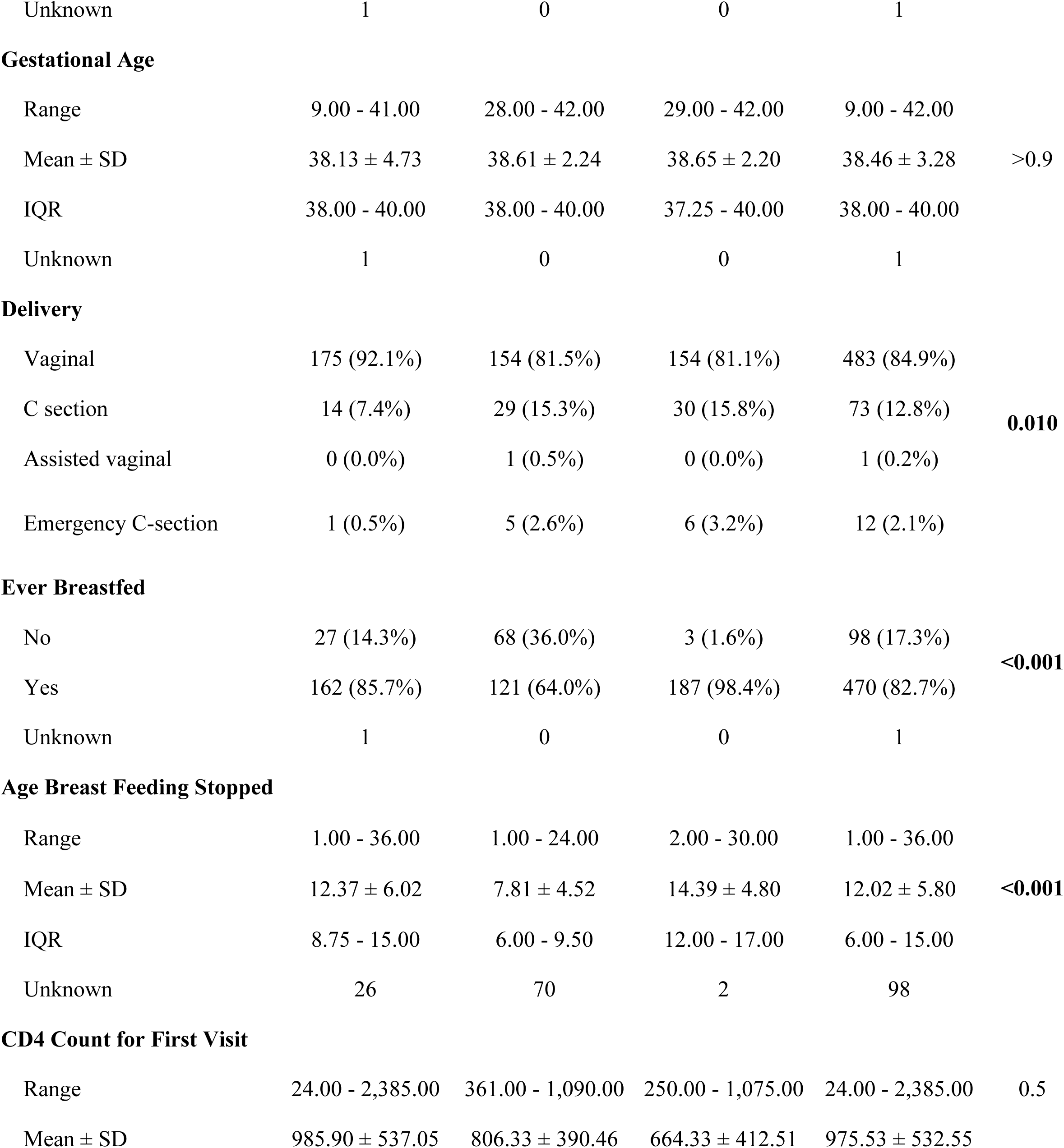
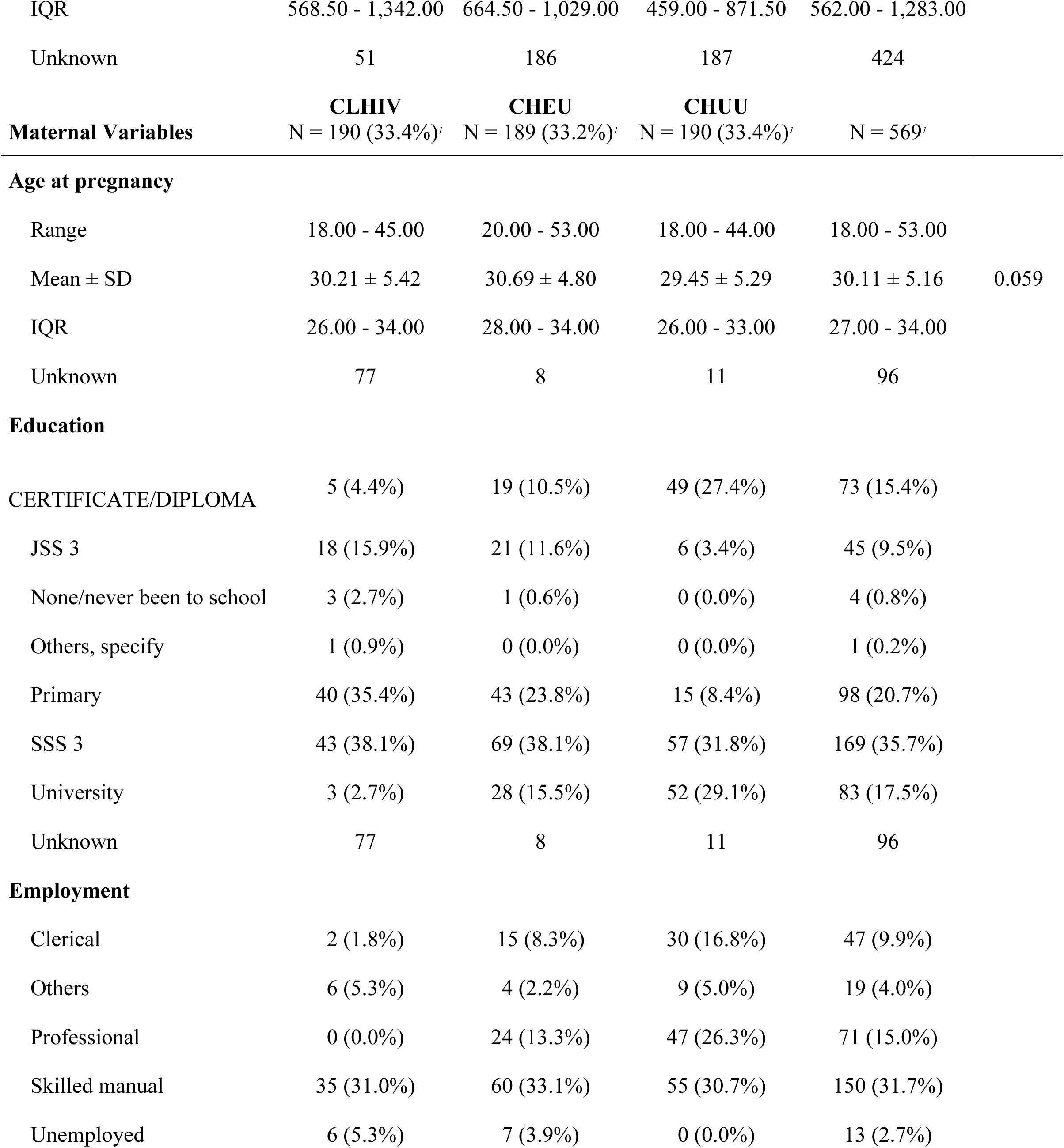
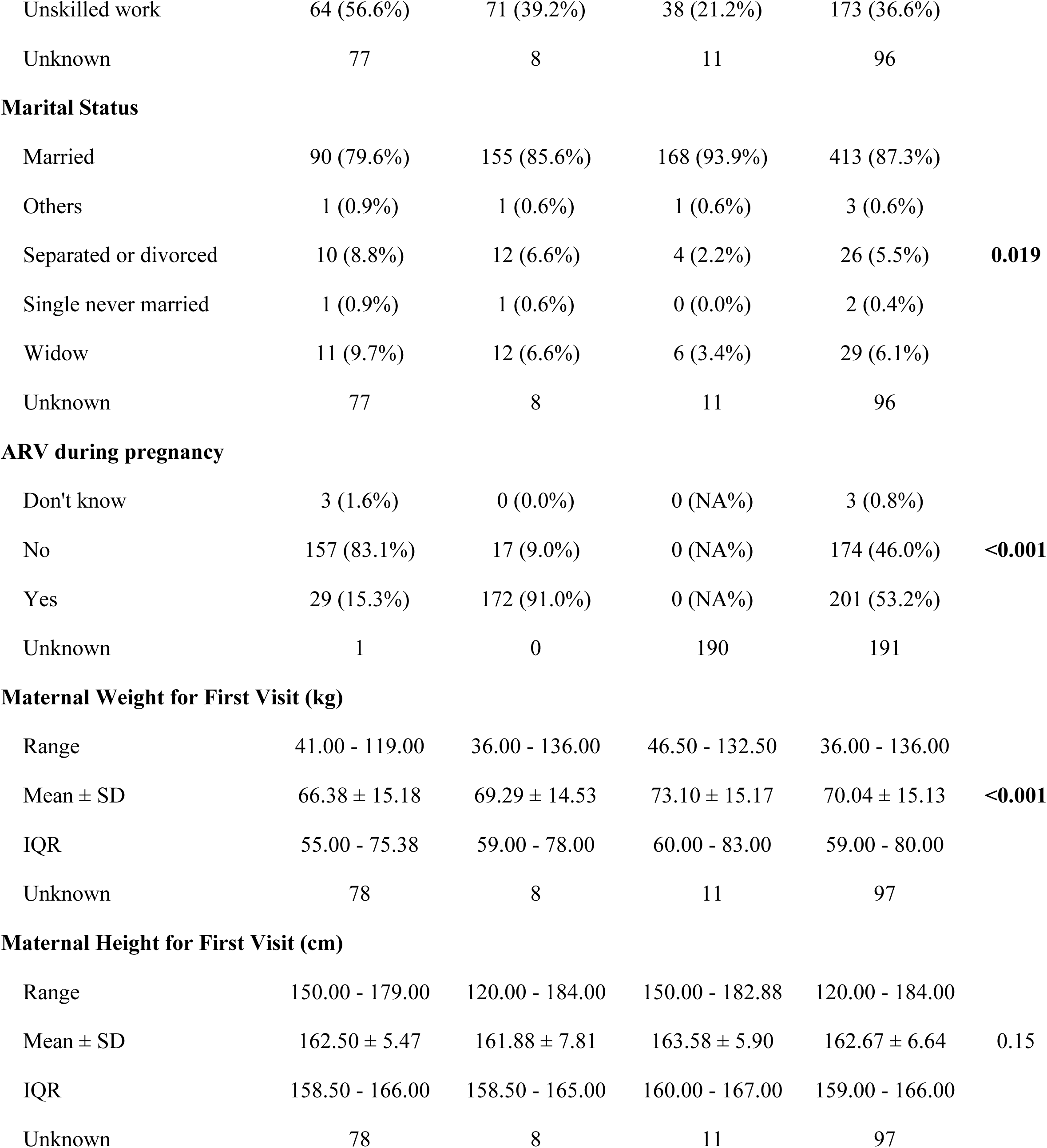
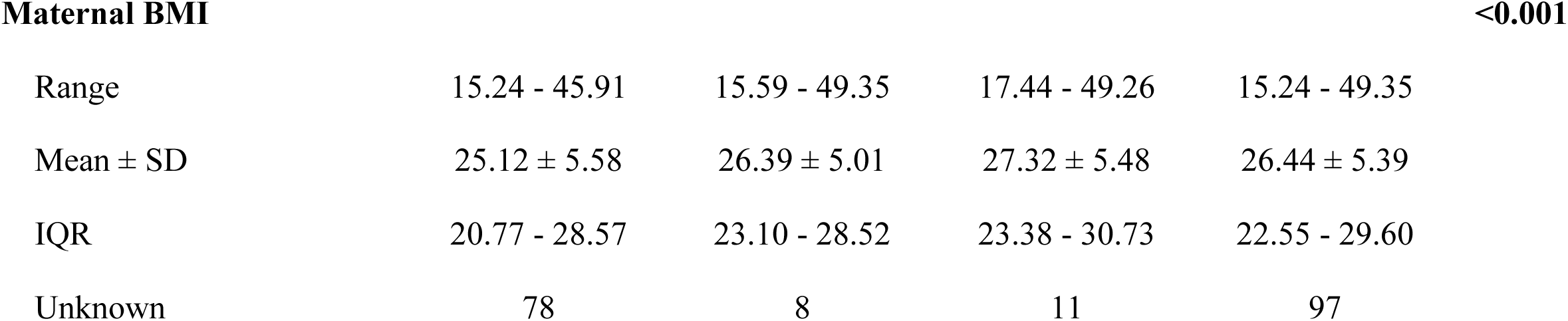
Baseline characteristics of children and mothers by HIV exposure and infection status. Values are presented as mean ± SD, median [IQR], or n (%) as appropriate. Comparisons were performed using Pearson’s Chi-squared test, Kruskal-Wallis rank-sum test, or Fisher’s exact test. Children living with HIV = CLHIV, Children HIV-exposed but uninfected = CHEU, Children HIV-unexposed and-uninfected (CHUU).

During the initial visit, weight and height were collected. CLHIV (21.32 ± 5.37 kg; 117.48 ± 13.64 cm) on average weighed less and were shorter in stature than CHEU (22.82 ± 7.26 kg; 22.82 ± 7.26 cm) and CHUU (24.62 ± 7.06 kg; 24.62 ± 7.06 cm) children. CHEU were shorter and lighter than CHUU. At baseline, BMI was significantly different depending on the children’s exposure/infection status.

In examining maternal or perinatal characteristics from this study cohort, there were 4 different forms of delivery: vaginal, cesarean, assisted vaginal, and emergency cesarean. Vaginal birth was most common in all groups and significantly different across study groups (**Table 1**); however, breastfeeding duration differed significantly (p <0.001). CHUU were breastfed the longest (14.39 months), compared with CLHIV (13.37 months) and CHEU (7.81 months). Other maternal characteristics were also gathered during the initial visit. Mothers of CLHIV had the lowest mean weight (66.38 kg) and BMI (25.12 kg/m^2^), compared with mothers of CHEU (69.29 kg; 26.4 kg/m^2^) and CHUU (73.10 kg; 27.3 kg/m^2^) (**Table 1**). Mothers of CHEU weighed less on average than mothers of CLHIV, while maternal height did not differ significantly across CLHIV, CHEU, and HUU groups. 78 mothers of CLHIV were excluded from BMI analysis due to missing height or weight; therefore, BMI estimations could not be determined accurately for these cases.

**Figure 1** depicts the different z-scores (WAZ, HAZ, and BAZ) of the 3 study groups along with the duration of the study period. Across all visits, CLHIV had consistently lower WAZ and HAZ scores compared to CHEU and CHUU, with CHUU individuals having the highest z-score. While BAZ slowly decreases throughout the entirety of the study but decreases at a larger rate in 7.5 to 7.9-year-olds, up until 10, where it plateaus, and then it slowly begins to increase. CHEU and CHUU have slight decreases until approximately 6 years old, where CHUU will remain constant until eventually increasing around 9.5 years old for WAZ. After 6 years old, CHEU z-score decreases around 7.5 years of age.

**Figure 1A-C.**
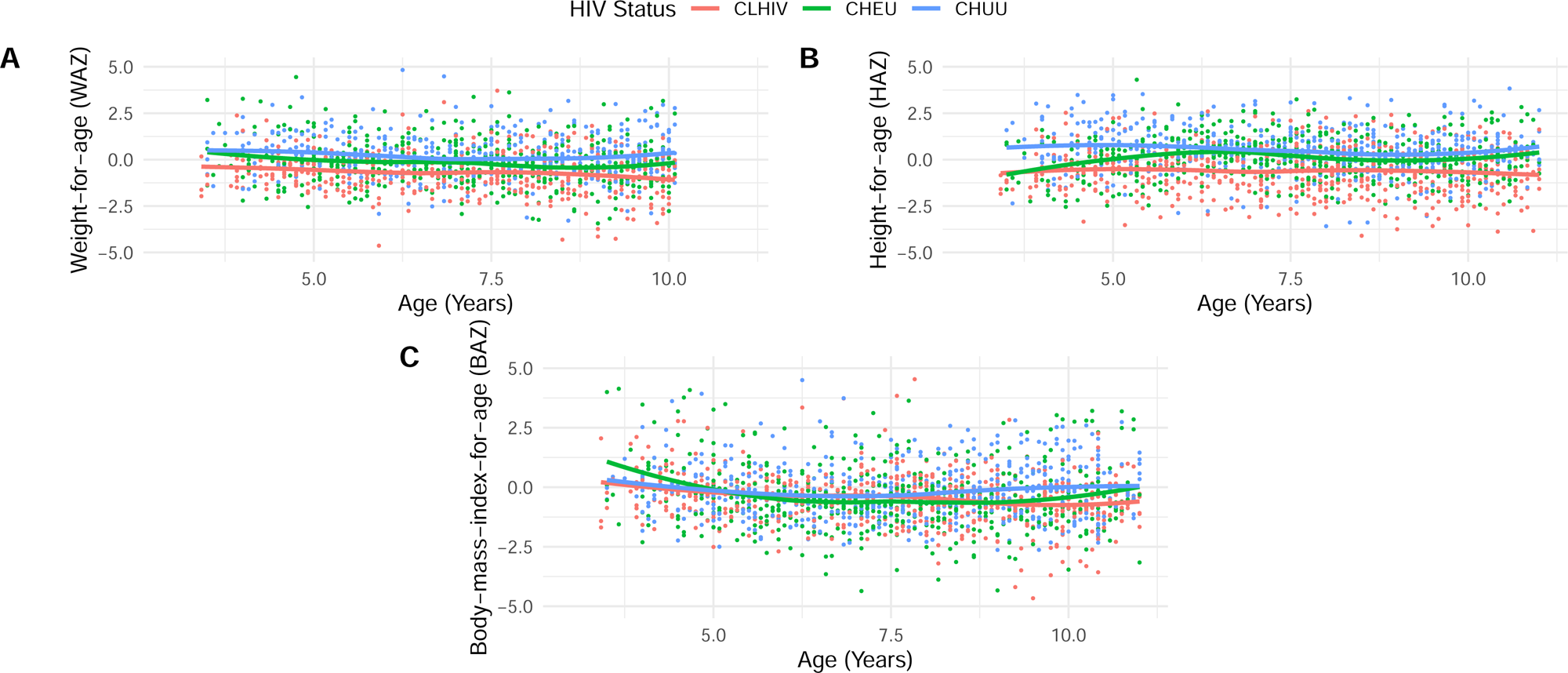
Anthropometric growth Z-scores among Children living with HIV (CLHIV), children HIV-exposed but uninfected (CHEU), children HIV-unexposed and-uninfected (CHUU) aged 3-11 years. (A) Weight-for-age (WAZ), (B) Height-for-age (HAZ), and (C) Body-mass-index-for-age (BAZ) Z-scores by age and HIV group. Lines represent locally weighted regression (LOESS) smoothed means with 95% confidence intervals. CLHIV consistently show lower Z-scores across all indicators compared with CHEU and CHUU

### Antiretroviral (ARV) related data for CLHIV

**Table 2** presents Antiretroviral (ARV) related data for CLHIV. On average, these children started ARV at a mean age of 37 months (SD 30.2, Range 41-126 months). At baseline, the median CD4 count was 986 cells/mm^3^ (IQR 568 - 1342). Although viral load data were collected, a large portion of values were missing or undetectable (120/190), were therefore excluded from analysis.

**Table 2.**
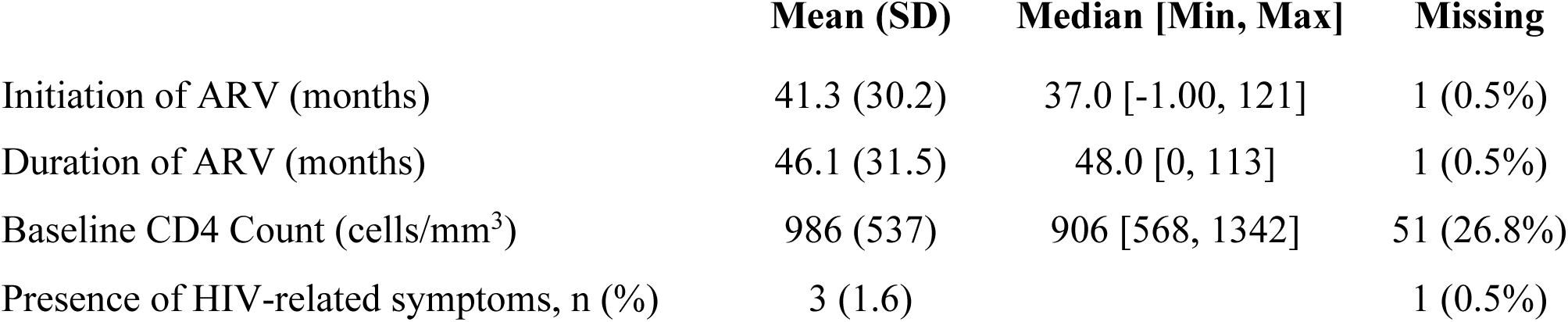
Antiretroviral therapy (ARV) characteristics and baseline immunologic markers among HIV-infected (HI) children. Values are presented as mean ± SD or median [IQR]. Viral load data were excluded due to high rates of missing or undetectable values. CD4 count = cluster of differentiation 4 lymphocyte count (cells/mm^3^); ARV = antiretroviral therapy

### Descriptive Statistics and Prevalence of Malnutrition

Descriptive statistics are presented in **Table 3A** for WAZ, HAZ, and BAZ throughout the three different visitations. Visit 1 is considered the baseline. Across visits, CLHIV consistently had lower WAZ and HAZ scores than CHEU and CHUU; CHUU had the highest z-score (**Table 3A**). CHEU and CHUU improved after the first visit, while CLHIV became stagnant after the second.

**Table 3.** Descriptive statistics and prevalence of malnutrition across visits by HIV status. Values represent Mean ± SD (IQR) unless otherwise noted or n (%). **WFH excluded due to >65% missingness (387 observations).** WAZ= Weight-for-Age Z-score; HAZ= Height-for-Age Z-score; BAZ= Body-Mass-Index-for-Age Z-score.

**A. Anthropometric Z-scores by Visit and HIV Status**
| Indicator | Visit | CLHIV | CHEU | CHUU | p-value |
| --- | --- | --- | --- | --- | --- |
| **WAZ** | Visit 1 | $-0.89 \pm 1.37$ (−1.49, −0.19) | $-0.32 \pm 1.19$ (−1.12, 0.33) | $0.15 \pm 1.11$ (−0.57, 0.73) | **< 0.001** |
| | Visit 2 | $-0.68 \pm 1.05$ (−1.27, −0.06) | $-0.15 \pm 1.19$ (−0.87, 0.43) | $0.23 \pm 1.14$ (−0.36, 0.91) | **0.027** |
| | Visit 3 | $-0.71 \pm 1.38$ (−1.21, 0.09) | $-0.13 \pm 1.18$ (−0.90, 0.46) | $0.22 \pm 1.09$ (−0.49, 0.78) | **0.032** |
|  | Missing (n) | 13 / 29 / 47 | 7 / 19 / 41 | 5 / 22 / 34 |  |
| **HAZ** | Visit 1 | $-1.05 \pm 2.22$ (−1.67, 0.00) | $-0.19 \pm 1.42$ (−0.83, 0.60) | $0.25 \pm 1.37$ (−0.48, 1.04) | **< 0.001** |
| | Visit 2 | $-0.64 \pm 1.34$ (−1.44, 0.22) | $0.16 \pm 1.08$ (−0.56, 0.80) | $0.57 \pm 1.12$ (0.00, 1.26) | **< 0.001** |
| | Visit 3 | $-0.65 \pm 2.17$ (−1.36, 0.32) | $0.31 \pm 1.06$ (−0.35, 1.00) | $0.67 \pm 1.10$ (0.00, 1.26) | **< 0.001** |
|  | Missing (n) | 0 / 0 / 0 | 0 / 0 / 0 | 0 / 0 / 0 |  |
| **BAZ** | Visit 1 | $-0.43 \pm 1.39$ (−1.24, 0.29) | $-0.31 \pm 1.77$ (−1.21, 0.27) | $0.00 \pm 1.32$ (−0.89, 0.62) | **0.020** |
| | Visit 2 | $-0.49 \pm 0.99$ (−1.08, 0.11) | $-0.38 \pm 1.34$ (−1.11, 0.07) | $-0.15 \pm 1.21$ (−1.10, 0.53) | 0.13 |
| | Visit 3 | $-0.54 \pm 0.95$ (−1.06, 0.02) | $-0.49 \pm 1.33$ (−1.30, 0.02) | $-0.23 \pm 1.37$ (−1.10, 0.42) | 0.30 |
|  | Missing (n) | 1 / 0 / 1 | 0 / 0 / 0 | 0 / 0 / 0 |  |

| Variable | CLHIV | CHEU | CHUU | p-value<sup>2</sup> |
| --- | --- | --- | --- | --- |
| *Visit 1* | *N = 190 (33.4%)<sup>1</sup>* | *N = 189 (33.2%)<sup>1</sup>* | *N = 190 (33.4%)<sup>1</sup>* |  |
| **Stunting** |  |  |  |  |
| Non-Stunted | 154 (81.1%) | 179 (94.7%) | 181 (95.3%) | **<0.001** |
| Stunted | 36 (18.9%) | 10 (5.3%) | 9 (4.7%) |  |
| **Underweight** |  |  |  |  |
| Non-Underweight | 157 (88.7%) | 175 (96.2%) | 182 (98.4%) | **<0.001** |
| Underweight | 20 (11.3%) | 7 (3.8%) | 3 (1.6%) |  |
| Unknown | 13 | 7 | 5 |  |
| **Wasting** |  |  |  |  |
| Non-wasting | 175 (92.6%) | 173 (91.5%) | 186 (97.9%) | **0.020** |
| Wasting | 14 (7.4%) | 16 (8.5%) | 4 (2.1%) |  |
| Unknown | 1 | 0 | 0 |  |
| **Malnourished** |  |  |  |  |
| Normal | 124 (68.5%) | 118 (64.8%) | 144 (75.8%) | 0.2 |
| Mild Malnutrition | 49 (27.1%) | 53 (29.1%) | 41 (21.6%) |  |
| Moderate Malnutrition | 8 (4.4%) | 11 (6.0%) | 5 (2.6%) |  |
| Unknown | 9 | 7 | 0 |  |
| *Visit 2* | *N = 183 (33.2%)<sup>1</sup>* | *N = 185 (33.5%)<sup>1</sup>* | *N = 184 (33.3%)<sup>1</sup>* |  |
| **Stunting** |  |  |  |  |
| Non-stunted | 159 (86.9%) | 181 (97.8%) | 180 (97.8%) | **<0.001** |
| Stunted | 24 (13.1%) | 4 (2.2%) | 4 (2.2%) |  |
| **Underweight** |  |  |  |  |
| Non-underweight | 141 (91.6%) | 158 (95.2%) | 159 (98.1%) | **0.027** |
| Underweight | 13 (8.4%) | 8 (4.8%) | 3 (1.9%) |  |
| Unknown | 29 | 19 | 22 |  |
| **Wasting** |  |  |  |  |
| Non-wasting | 171 (93.4%) | 172 (93.0%) | 179 (97.3%) | 0.13 |
| Wasting | 12 (6.6%) | 13 (7.0%) | 5 (2.7%) |  |
| **Malnourished** |  |  |  |  |
| Normal | 125 (69.1%) | 121 (66.5%) | 130 (70.7%) | 0.8 |
| Mild Malnutrition | 45 (24.9%) | 48 (26.4%) | 46 (25.0%) |  |
| Moderate Malnutrition | 11 (6.1%) | 13 (7.1%) | 8 (4.3%) |  |
| Unknown | 2 | 3 | 0 |  |
| *Visit 3* | *N = 176 (33.5%)<sup>1</sup>* | *N = 178 (33.9%)<sup>1</sup>* | *N = 171 (32.6%)<sup>1</sup>* |  |
| **Stunting** |  |  |  |  |
| Non-stunted | 154 (87.5%) | 175 (98.3%) | 168 (98.2%) | **<0.001** |
| Stunted | 22 (12.5%) | 3 (1.7%) | 3 (1.8%) |  |
| **Underweight** |  |  |  |  |
| Non-underweight | 118 (91.5%) | 129 (94.2%) | 135 (98.5%) | **0.032** |
| Underweight | 11 (8.5%) | 8 (5.8%) | 2 (1.5%) |  |
| Unknown | 47 | 41 | 34 |  |
| **Wasting** |  |  |  |  |
| Non-wasting | 164 (93.7%) | 164 (92.1%) | 164 (95.9%) | 0.3 |
| Wasting | 11 (6.3%) | 14 (7.9%) | 7 (4.1%) |  |
| Unknown | 1 | 0 | 0 |  |
| **Malnourished** |  |  |  |  |
| Normal | 116 (66.7%) | 106 (60.6%) | 117 (68.4%) | 0.6 |
| Mild Malnutrition | 45 (25.9%) | 52 (29.7%) | 43 (25.1%) |  |
| Moderate Malnutrition | 13 (7.5%) | 17 (9.7%) | 11 (6.4%) |  |
| Unknown | 2 | 3 | 0 |  |

<sup>1</sup>n (%)
<sup>2</sup>Pearson's Chi-squared test; Fisher's exact test

To examine growth patterns within the three study groups, groups were classified based on three factors in categorical fashion (yes/no): Stunting (HAZ ≤ -2), Underweight (WAZ ≤ -2), Wasting (BAZ ≤ -2) (**Table 3B**). We also examined malnutrition based on BAZ scores alone: Normal (BAZ > 1), Mild (BAZ = -1 to -1.9), and Moderate (BAZ = -2.0 to -3) (**Table 3B**).

Across the three visitations, CLHIV were more likely to experience stunting (36 children, 18.9%), although this number steadily reduced at later visits (12.5% by visit 3), CHEU (1.7%), and CHUU (1.8%) had a lower stunting prevalence and greater recovery (p <0.001).

### Cohort-Time Effect

Of the 569 children enrolled, 92% completed all three visits. Dropout analysis revealed no significant baseline difference in age, sex, HIV status, maternal BMI, delivery method, or anthropometric Z-scores between retained and lost participants (all p>0.1). However, dropouts were less likely to have been breastfed (77% vs 85%, p = 0.03).

A generalized estimating equation (GEE) analysis was done for the different z-scores (WAZ, HAZ, and BAZ adjusted for HIV status, visit number, maternal BMI, gender, whether they were breastfed, and delivery method) (**Table 4**). Dietary intake and maternal employment status were explored but excluded due to poor model fit (high QIC). Although egg consumption was significant in preliminary cross-sectional models, dietary models were excluded from GEE due to poor model fit. Final analyses are presented using the more efficient and simpler parsimonious model, which excludes diet and employment.

**Table 4.** Adjusted generalized estimating equation (GEE) models for longitudinal growth outcomes. Estimates (β) with 95% confidence intervals (CI) and p-values are presented. Models were adjusted for visit number, maternal BMI, child sex, breastfeeding status, and delivery method. Bold values indicate statistical significance (p <0.05).

| *Predictors* | **WAZ** |  | **HAZ** |  | **BAZ** |  |
| --- | --- | --- | --- | --- | --- | --- |
| | $\beta$ (95 % CI) | p | $\beta$ (95 % CI) | p | $\beta$ (95 % CI) | p |
| (Intercept) | -1.09 (-1.51 – -0.68) | **0.001** | -0.44 (-0.88 – -0.00) | 0.190 | -1.37 (-1.84 – -0.91) | **<0.001** |
| Reference: CLHIV | -0.67 (-0.84 – -0.49) | **<0.001** | -1.04 (-1.22 – -0.86) | **<0.001** | -0.13 (-0.32 – 0.05) | 0.302 |
| Reference: CHEU | -0.35 (-0.50 – -0.19) | **0.009** | -0.38 (-0.55 – -0.22) | **0.003** | -0.17 (-0.35 – 0.01) | 0.230 |
| Visit: Visit 2 | 0.12 (-0.03 – 0.27) | **<0.001** | 0.32 (0.16 – 0.48) | **<0.001** | -0.11 (-0.28 – 0.06) | **0.047** |
| Visit: Visit 3 | 0.13 (-0.03 – 0.28) | **0.001** | 0.46 (0.29 – 0.62) | **<0.001** | -0.19 (-0.37 – -0.02) | **0.002** |
| Maternal BMI | 0.04 (0.03 – 0.06) | **<0.001** | 0.02 (0.01 – 0.04) | **0.016** | 0.05 (0.03 – 0.06) | **<0.001** |
| Sex: Female | -0.07 (-0.20 – 0.05) | 0.481 | 0.08 (-0.05 – 0.22) | 0.444 | -0.17 (-0.31 – -0.03) | 0.115 |
| Breast Fed: YES | 0.03 (-0.15 – 0.22) | 0.833 | 0.05 (-0.14 – 0.24) | 0.758 | 0.08 (-0.12 – 0.28) | 0.636 |
| Delivery Method |  |  |  |  |  |  |
| C Section | 0.14 (-0.04 – 0.33) | 0.334 | -0.04 (-0.23 – 0.15) | 0.751 | 0.31 (0.11 – 0.51) | 0.051 |
| Assisted Vaginal | -0.23 (-1.50 – 1.04) | 0.111 | -0.02 (-1.44 – 1.41) | 0.900 | -0.39 (-1.90 – 1.13) | **0.011** |
| Emergency C Section | -0.25 (-0.66 – 0.17) | 0.486 | -0.01 (-0.46 – 0.45) | 0.981 | -0.44 (-0.92 – 0.05) | 0.305 |
| N | 450<sub>ID</sub> |  | 472<sub>ID</sub> |  | 472<sub>ID</sub> |  |
| Observations | 1210 |  | 1394 |  | 1394 |  |

When controlling for these factors, the study timepoint (visits) and HIV status were significant for WAZ (p = 0.002) and BAZ (p < 0.001). WAZ increased slightly at visit 2-3 (Visit 2: β = 0.12, 95% CI: -0.03 – 0.27; Visit 3: β = 0.13, 95% CI: -0.03 - 0.28), because the 95% confidence intervals cross and include zero, which indicates no statistical significance. HAZ increased significantly at visit 2 (β = 0.32; p < 0.001) and at visit 3 (β = 0.46; p < 0.001). BAZ slightly declined by visit 3 (β =-0.19, p = 0.002).

HIV status remained significant for both WAZ and HAZ, but not BAZ. Compared to CHUU, CLHIV had lower WAZ (β = -0.67; p < 0.001) and HAZ (β = -1.04; p < 0.001), while CHEU had intermediate deficits (WAZ: β = -0.35, p = 0.009; HAZ: β = -0.38, p = 0.003). Maternal BMI was positively associated with all growth outcomes (WAZ: β =0.04, p<0.00; HAZ: β =0.02, p=0.016; BAZ β = 0.05, p<0.001). The interaction effects of gender, breastfeeding, and delivery methods were not statistically significant.

We performed a GEE analysis of the interaction between dietary frequency and HIV status. As depicted in **Supplemental Figure 1A-D**, suggested potential modifying effects for BAZ were observed, but these models had higher QIC values and were not retained in the final analysis. Similarly, an exploratory linear regression models were performed on the different Z scores, indicating that higher egg consumption was associated with improved WAZ. **Supplemental Table 1** is a table that summarizes the coefficients, 95% CI, and p-values for associations between dietary frequency variables (eggs, beans, meat, vegetables, and multivitamins) and WAZ in both linear regression and GEE. The model was then evaluated using QIC for the GEE models. Although egg consumption was significant in cross-sectional analyses, this association did not remain significant in the longitudinal GEE analysis.

## Discussion

This study is among the first to examine the impact of perinatal HIV infection and exposure on longitudinal growth among school-aged children in Nigeria, comparing CLHIV, CHEU, and CHUU. Our study showed that even in the context of universal maternal ART and breastfeeding, CLHIV had consistently lower weight-for-age (WAZ) and height-for-age (HAZ) z-scores, indicating persistent deficiencies in both linear and ponderal growth despite ART access^25^. CHEU experienced intermediate growth outcomes, better than CLHIV but still lower than CHUU, which suggests partial recovery but not normalization^3,17,20^. These findings confirm that perinatal HIV infection continues to impair physical growth into school age, consistent with recent global analysis of HIV-exposed uninfected communities^16^.

As in prior studies ^7,18,26–28^, CLHIV showed lower HAZ and BAZ scores than both CHEU and CHUU, indicating a higher burden of stunting. CLHIV persistently showed inadequate linear growth despite ART access and successful viral suppression^11,28,29^. While perinatal HIV exposure/infection is a key determinant, early childhood development and environmental factors are also crucial in influencing growth potential. CLHIV often experience chronically increased resting energy expenditure (REE) and inefficient energy utilization^25,29–31^, typically due to recurrent infections, persistent immune activation, and altered metabolic signaling^12,32–34^. These elevated energetic demands divert calories away from linear growth and tissue accretion, making growth catch-up difficult despite the presence of ART and adequate caloric intake^17,20,29^.

Early deficits in bone mineral accrual are driven by inflammation, reduced physical activity, micronutrient deficiencies, and ART-reduced mitochondrial effects, may impair skeletal growth trajectories and limit height catch-up capacities ^13,25,35,36^. These metabolic, environmental, and developmental constraints together help explain persistent stunting and underweight observations seen across CLHIV despite viral suppression and ART exposure.

Among CLHIV, those with later initiation to Antiretroviral therapy (ART) had lower WAZ, HAZ, and BAZ scores and were more likely to be stunted or underweight in CLHIV. In this cohort, ART initiation occurred at a mean age of approximately 41 months. Consistent with previous studies, early ART initiation shows a positive correlation and better anthropometric z-scores^18,29,37^.

Although CHEU are not HIV infected, their perinatal exposure to HIV and ART likely disrupts immune maturation, increases inflammatory tone, and alters metabolic and gastrointestinal pathways^8,14,16,32^. Prior studies have shown that CHEU exhibit higher rates of early infections, immune activation, and subtle placental insufficiency, along with mitochondrial alterations linked to maternal ART exposure^8,22^. These factors increase energetic demand, impaired energy metabolism and increased oxidative stress, while reducing the efficiency of nutrient utilization^12,14^. CHEU have documented increased risks of impaired antibody transfer, greater healthcare utilization, and early deficits in growth and neurodevelopment^38^. When combined with maternal health and socioeconomic constraints, these biological factors may contribute to persistent anthropometric deficits and limit recovery despite the absence of HIV infection^35,39^.

Among early childhood developmental factors that play a role in malnutrition in this study is socioeconomic status (SES). SES is measured by maternal demographics: education, employment, and marital status. It is observed that throughout the three study groups, CHEU and CHUU came from mothers with a higher educational background when compared to those of CLHIV. Mothers with a higher education are more likely to have CHUU, while those with a disadvantaged SES are more likely to have CLHIV^39^. Of 83 mothers who attended university, 29.1% CHUU, 15.5% CHEU, and only 2.7% were CLHIV. Education status has been shown to have a positive correlation to HIV infection status among maternal and child status^1^. Education likely influences access and utilization of essential health resources for CLHIV to better maintain HIV, and the same goes for those who are CHEU. SES can be reflected by maternal careers, and education may be a direct indicator of which careers that are available to pursue. CHEU and CHUU mothers are more likely to hold skilled manual (60 and 55, respectively) and professional careers (24 and 47, respectively). While 56.6% mothers of CLHIV are more likely to do unskilled work.

Potential drivers of lower growth outcomes, such as maternal BMI, lower maternal BMI, and poor environmental conditions, were linked to higher odds of stunting^35^. Prior studies^29,40^ have shown that maternal malnutrition is commonly associated with childhood malnutrition, suggesting stunting may be intergenerational. Women who were stunted in childhood are more likely to have offspring who are stunted^32^, indicating the persistent impact of early-life maternal nutrition on subsequent generations. Maternal malnutrition during the gestational period can also decrease fetal growth, causing lower birth weight and impaired postnatal development.

This study examined the long-term implications of poor growth outcomes in children. Those exhibiting signs of malnutrition (stunting, wasting, and/or underweight) are at a greater risk for experiencing failure to thrive, reduction in resting energy, impaired cognitive development, and hypertension, which can lead to an increase in morbidity and mortality^33,36,41^. Impaired linear growth also adversely affects the quality of life by decreasing/limiting physical capacity. Without early intervention, these growth deficits can have enduring consequences throughout childhood and beyond^42^. Although diet is a known determinant in growth outcome, it was omitted from the final models’ due lack of significance. Consequently, findings did not demonstrate a meaningful correlation between dietary frequency and anthropometric Z-scores within this cohort.

A significant strength of this study is the simultaneous examination of all three study groups - CLHIV, CHEU, and CHUU age- and sex-matched within a cohort study. This design allows for direct comparison of both HIV infection and exposure effects on growth outcomes, whereas many prior studies compare only two groups or lack appropriate unexposed controls. Focusing on school-aged children was a unique strength, given that prior studies in Nigeria and SSA have focused on infancy/early childhood or adolescence, rarely including CLHIV as a study group. By including all three groups, this study allows for a more nuanced understanding of how perinatal HIV exposure and infection differentially impact growth.

There are, however, some limitations to note. While a longitudinal analysis, only three visits were available for this analysis, given the allotted duration of follow-up. Other studies with similar methodologies followed a longer duration period, 18-24 months, and had more frequent visits on a quarterly/monthly basis. Although overall dropout rates were low (8%), children who dropped out were less likely to be breastfed. Differences in breastfeeding status between retained and dropout participants may limit the generalizability of findings related to breastfeeding, and results should be interpreted with this in mind. WFH did not apply to most of the cohort’s age range; therefore, it could not be longitudinally analyzed, but BAZ was then used as the indicator for wasting as per WHO guidance^43^. Another limitation is data unavailability – for example, birth metrics were not available; birth metrics (birth length) could have helped lead to some prediction of the child’s later height. Also, birth length could have given some insight into the child’s future weight, excluding those that may have been born prematurely. Collecting data on vitamin deficiency and diarrhea frequency could have provided additional insights into factors contributing to stunting within this population. Studies have shown that children who are more susceptible to diarrhea and other infections are more likely to experience worsening malnutrition, and early-life viral co-infections such as cytomegalovirus have also been linked to growth impairments and immune activation among HIV-exposed^34,44^. Viral load data were also incomplete due to a high proportion of undetectable or missing values, which limited the ability to assess virological suppression in relation to growth outcomes. Furthermore, maternal SES and its alignment with children’s HIV status highlighted some limitations. Although not implicitly selected this way, data express that CLHIV and CHEU tend to come from socioeconomic groups of the lower class when compared to those of CHUU. The lack of variety aligns with CLHIV and CHEU often coming from lower SES groups than CHUU^20^. However, we were unable to fully control for SES given the disparate determinants of health among the groups. Despite attempts to match participants through community referrals, CLHIV and CHEU were disproportionately from lower SES backgrounds compared to CHUU, reflecting the well-documented association between HIV and poverty in this setting.

In conclusion, we found an association between HIV exposure/infection with childhood growth outcomes (WAZ, HAZ, and BAZ) among school-aged children in Nigeria, suggesting that growth deficits of CLHIV and CHEU remain prevalent after birth and extend into childhood.

Future research should focus on this relationship to examine and better understand the consequences on CLHIV, as well as understanding how interventions might improve growth outcomes and development. In this study, understanding the importance of evaluating CHEU due to their scores also being impacted when compared to CHUU. Although their growth outcomes were not as low as those of CLHIV, they should still be examined. Collecting information on nutrition/diet, access to clean water, maternal health and education, and overall gut health should also be studied due to their influence on childhood growth outcomes. Children who are born to mothers who are HIV infected, whether the child is affected or uninfected, need special attention throughout and beyond their adolescence. If not given, they run a high risk of detrimental and long-term impairments in their health, development, and growth. A clear understanding of these impairments can support the development of strategies to mitigate and control HIV-linked growth deficiencies.

## Data Availability

The data that supportt the findings of this study are available from the corresponding author upon resonable request, subject to institutional review and ethical approval to ensure participant confidentiality.

## Acknowledgments

The authors are grateful to the children and families who made this study possible and to all the study staff of the University of Benin Teaching Hospital and Institute of Human Virology, Nigeria. Special thanks to all members of the DOMHaIN Study Team, which comprises the authors and the following team members: Mr. Oghenenero Igedegbe, Mr. Ruxton Adebiyi, Matron Christy Ndekwu, Dr. Uwagboe Odigie, Ms. Oyemwen Olaye, Ms. Ehioze Awanlemhen, Mr. Samuel Chukwumaeze, Mr. Matthew Imoe, Mr. Daniel Oakhu, and Ms. Susan Dare are acknowledged for the recruitment, sample, and data collection. The authors thank Dr. Nosakhare Idemudia, Mr. Osasumwen Ehigie, Mr. Kelly Avenbuan, and Ms. Amara Godwins for laboratory management and support with sample processing. Drs. Nneka Chukwumah Stanley Iyorzor, Owen Omorogbe, and Chioma Ugorji are acknowledged for the clinical examination during study visits and for their flexibility with recruitment and scheduling.

## Funding

This work was partly supported by a grant from the National Institutes of Health (NIDCR R01DE028154 and R01DE032216). The funder had no role in the study design, collection, analysis, interpretation of the data, or manuscript writing.

## Conflict of Interest

No competing interest

## Ethics approval and consent to participate

Before the commencement of the study, ethical approvals from the Institutional Review Boards at UBTH (ADM/E22/AVOL.VII/14713, 31 January 2019), University of Maryland, Baltimore (HP-00084081), and Rutgers State University of New Jersey (Pro2019002047) were obtained. All procedures performed in studies involving human participants were in accordance with the tenets of the Declaration of Helsinki. All the study activities, benefits/risks of voluntary participation, and withdrawal from the study were verbally communicated to parents/guardians or caregivers in English and/or pidgin English. Questions were asked to confirm that they understood the study, and written informed consent was obtained before recruitment.

---

**Figure 1A-D.** Anthropometric growth Z-scores among Children living with HIV (CLHIV), children HIV-exposed but uninfected (CHEU), children HIV-unexposed and-uninfected (CHUU).

**Supplemental Figure 1A-D.** Exploratory GEE models assessing the interaction between dietary frequency and HIV status on body-mass-index-for-age (BAZ). As noted in the methods, Weight-for-height (WFH) was not included in the inferential analyses but is represented here for completeness. Estimated marginal means (±95% CI) are shown across diet frequency categories (rarely, weekly, daily) for HI, HEU, and HUU. Although diet-by-HIV interactions reached significance in some categories, overall model fit (QIC) was inferior to that of the model retained in the final analysis.

**Supplemental Table 1. Comparison of cross-sectional linear regression and longitudinal GEE models for dietary frequency predictors of weight-for-age Z-scores (WAZ).** Models include eggs, beans, meats, vegetables, and multivitamins as frequency variables. Results are shown as regression coefficients (β), 95% CI, and p-values. Significant results are bolded (p <0.05). The GEE model demonstrated better model fit (QIC = 261) than cross-sectional analyses.

## Abbreviations

ART: Antiretroviral therapy
DOMHaIN: Dental caries and its association with oral microbiomes and HIV in young children—Nigeria
HIV: Human immunodeficiency virus
CLHIV: Children Living with HIV
CHEU: Children HIV-exposed-but-uninfected
CHUU: Children HIV-unexposed-uninfected
SSA: sub-Saharan Africa

## Notes

### Competing Interest Statement

The authors have declared no competing interest.

### Author Declarations

The institutional Review Board of the University of Benin Teaching Hospital, the Institutional Review Board of the University of Maryland, Baltimore, and the Institutional Review Board of Rutgers State University of New Jersey gave ethical approval for this work. Written informed consent was obtained from parents or guardians of all participants prior to enrollment.

## References

1. Bassey, A. E. & Miteu, G. D. A review of current trends in HIV epidemiology, surveillance, and control in Nigeria. Annals of Medicine & Surgery 85, 1790–1795 (2023).

2. Awofala, A. A. & Ogundele, O. E. HIV epidemiology in Nigeria. Saudi Journal of Biological Sciences 25, 697–703 (2018).

3. Carlqvist, A. et al. Comparative growth of children who are HIV-exposed and uninfected with those who are HIV-unexposed and uninfected at age 1 and 2 years in South Africa: a prospective cohort study. eClinicalMedicine 89, 103515 (2025).

4. Modjadji, P., Mokgalaboni, K., Phoswa, W. N., Mothiba, T. M. & Lebelo, S. L. Growth Patterns of HIV-Exposed and -Unexposed Infants in African Countries: A Systematic Review and Meta-Analysis. Children 12, 624 (2025).

5. Ekali, G. L. et al. Growth of children who are HIV-exposed but uninfected: a systematic review and meta-analysis. The Lancet Child & Adolescent Health 9, 234–247 (2025).

6. Lartey, A. et al. Maternal HIV is associated with reduced growth in the first year of life among infants in the Eastern region of Ghana: the Research to Improve Infant Nutrition and Growth (RIING) Project. Maternal & Child Nutrition 10, 604–616 (2014).

7. Abate, B. B., Aragie, T. G. & Tesfaw, G. Magnitude of underweight, wasting and stunting among HIV positive children in East Africa: A systematic review and meta-analysis. PLoS ONE 15, e0238403 (2020).

8. Ruck, C. E. & Smolen, K. K. Effect of Maternal HIV Infection on Infant Development and Outcomes. Front.Virol. 2, 885246 (2022).

9. López, M. et al. Risk of intrauterine growth restriction among HIV-infected pregnant women: a cohort study. Eur J Clin Microbiol Infect Dis 34, 223–230 (2015).

10. Mbewe, E. G. et al. Socioeconomic Status and Cognitive Function in Children With HIV: Evidence From the HIV-Associated Neurocognitive Disorders in Zambia (HANDZ) Study. JAIDS Journal of Acquired Immune Deficiency Syndromes 89, 56–63 (2022).

11. Jesson, J., et al. Stunting and growth velocity of adolescents with perinatally acquired HIV: differential evolution for males and females. A multiregional analysis from the IeDEA global paediatric collaboration. J Intern AIDS Soc 22, e25412 (2019).

12. Duggal, S., Chugh, T. D. & Duggal, A. K. HIV and Malnutrition: Effects on Immune System. Clinical and Developmental Immunology 2012, 1–8 (2012).

13. Black, R. E. et al. Maternal and child undernutrition and overweight in low-income and middle-income countries. The Lancet 382, 427–451 (2013).

14. Sudfeld, C. R. et al. Linear Growth Faltering Among HIV-Exposed Uninfected Children. JAIDS Journal of Acquired Immune Deficiency Syndromes 73, 182–189 (2016).

15. Akintan, P. E., Akinsulie, A., Temiye, E. & Esezobor, C. Prevalence of Wasting, Stunting, and Underweight Among HIV Infected Underfives’, in Lagos Using W.H.O z Score. Nig Q J Hosp Med 25, 124–128 (2015).

16. Prendergast, A. J. & Evans, C. Children who are HIV-exposed and uninfected: evidence for action. AIDS 37, 205–215 (2023).

17. Costa, P. R. F. et al. Growth and body composition of adolescents and young adults with perinatal HIV infection: a systematic review and meta-analysis. BMC Public Health 25, 717 (2025).

18. Mwambenu, B., Ramoloko, V., Laubscher, R. & Feucht, U. Growth and the pubertal growth spurt in South African adolescents living with perinatally-acquired HIV infection. PLoS ONE 17, e0262816 (2022).

19. Preterm birth. WHO https://www.who.int/news-room/fact-sheets/detail/preterm-birth.

20. Fowler, M. G. et al. Growth deficits in antiretroviral and HIV-exposed uninfected versus unexposed children in Malawi and Uganda persist through 60 months of age. AIDS 36, 573–582 (2022).

21. Ejigu, Y., Magnus, J. H., Sundby, J. & Magnus, M. C. Differences in Growth of HIV-exposed Uninfected Infants in Ethiopia According to Timing of In-utero Antiretroviral Therapy Exposure. Pediatric Infectious Disease Journal 39, 730–736 (2020).

22. Slogrove, A. L., Powis, K. M., Johnson, L. F., Stover, J. & Mahy, M. Estimates of the global population of children who are HIV-exposed and uninfected, 2000–18: a modelling study. The Lancet Global Health 8, e67–e75 (2020).

23. Coker, M. O. et al. Dental caries and its association with the oral microbiomes and HIV in young children—Nigeria (DOMHaIN): a cohort study. BMC Oral Health 21, 620 (2021).

24. Myatt, Mark, G. Ernest. zscorer: Child Anthropometry z-Score Calculator. (2019).

25. Chirindza, N. et al. Body composition, physical fitness and physical activity in Mozambican children and adolescents living with HIV. PLoS ONE 17, e0275963 (2022).

26. Blaauw, J. et al. The presence of enteropathy in HIV infected children on antiretroviral therapy in Malawi. PLoS ONE 19, e0298310 (2024).

27. Jumare, J. et al. Compromised Growth Among HIV-exposed Uninfected Compared With Unexposed Children in Nigeria. Pediatric Infectious Disease Journal 38, 280–286 (2019).

28. Traisathit, P. et al. Impact of antiretroviral treatment on height evolution of HIV infected children. BMC Pediatr 19, 287 (2019).

29. Masi-Leone, M. et al. Growth and Metabolic Changes After Antiretroviral Initiation in South African Children. Pediatric Infectious Disease Journal 40, 1004–1010 (2021).

30. Franco-Oliva, A. et al. High resting energy expenditure, less fat-free mass, and less muscle strength in HIV-infected children: a matched, cross-sectional study. Front. Nutr. 10, 1220013 (2023).

31. Musoke, P. M. et al. Growth, immune and viral responses in HIV infected African children receiving highly active antiretroviral therapy: a prospective cohort study. BMC Pediatr 10, 56 (2010).

32. Prendergast, A. J. & Humphrey, J. H. The stunting syndrome in developing countries. Paediatrics and International Child Health 34, 250–265 (2014).

33. Martins, V. J. B. et al. Long-Lasting Effects of Undernutrition. IJERPH 8, 1817–1846 (2011).

34. Ahmed, S. et al. Severity of Diarrhea and Malnutrition among Under Five-Year-Old Children in Rural Bangladesh. The American Journal of Tropical Medicine and Hygiene 89, 223–228 (2013).

35. Amaha, N. D. & Woldeamanuel, B. T. Maternal factors associated with moderate and severe stunting in Ethiopian children: analysis of some environmental factors based on 2016 demographic health survey. Nutr J 20, 18 (2021).

36. Jacobson, D. L. et al. Growth at 2 Years of Age in HIV-exposed Uninfected Children in the United States by Trimester of Maternal Antiretroviral Initiation. Pediatric Infectious Disease Journal 36, 189–197 (2017).

37. Ndongo, F. A. et al. Long-term outcomes of early initiated antiretroviral therapy in sub-Saharan children: a Cameroonian cohort study (ANRS-12140 Pediacam study, 2008–2013, Cameroon). BMC Pediatr 21, 189 (2021).

38. Ntuli, L. et al. Developmental challenges in infants who are HIV-exposed uninfected. Brain, Behavior, and Immunity 130, 106078 (2025).

39. Ogunmola, J. O., Oladosu, O. & Olamoyegun, A. M. Relationship between socioeconomic status and HIV infection in a rural tertiary health center. HIV 61 (2014) doi:10.2147/HIV.S59061.

40. Das, R. et al. Maternal Underweight and Its Association with Composite Index of Anthropometric Failure among Children under Two Years of Age with Diarrhea in Bangladesh. Nutrients 14, 1935 (2022).

41. Mabaya, L., Matarira, H. T., Tanyanyiwa, D. M., Musarurwa, C. & Mukwembi, J. Growth Trajectories of HIV Exposed and HIV Unexposed Infants. A Prospective Study in Gweru, Zimbabwe. Global Pediatric Health 8, 2333794X21990338 (2021).

42. Williams, P. L. & Jesson, J. Growth and pubertal development in HIV-infected adolescents. Current Opinion in HIV and AIDS 13, 179–186 (2018).

43. Weltgesundheitsorganisation. Length/Height-for-Age, Weight-for-Age, Weight-for-Length, Weight-for-Height and Body Mass Index-for-Age; Methods and Development. (WHO Press, Geneva, 2006).

44. Checkley, W. et al. Multi-country analysis of the effects of diarrhoea on childhood stunting. International Journal of Epidemiology 37, 816–830 (2008).

