## Supplementary figures and images for "Persistent Anthropometric Deficits in School-aged Children with Perinatal HIV Exposure"

### Supplemental Figure 1A-D

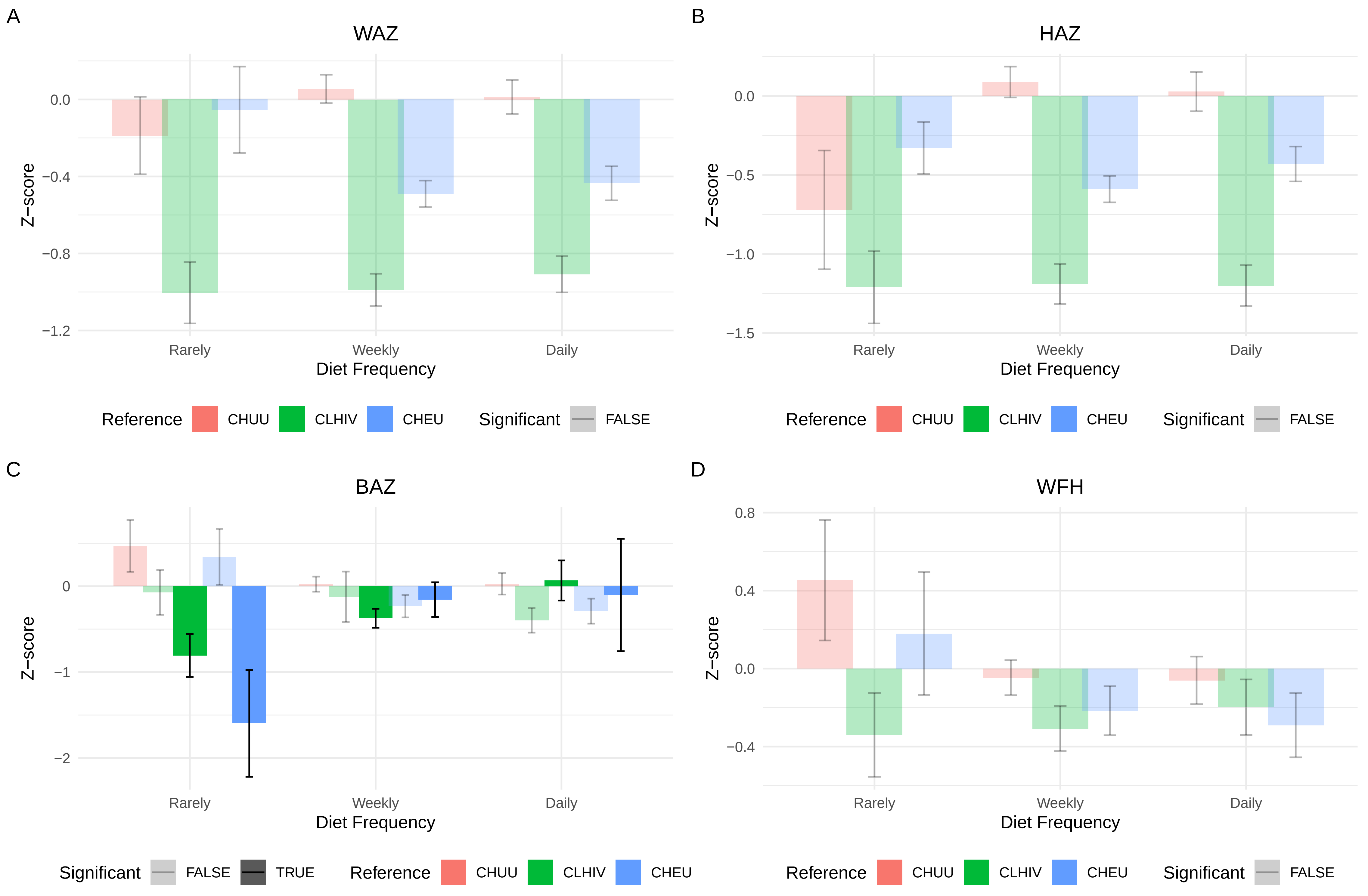
