## Supplemental Table 1 for "Persistent Anthropometric Deficits in School-aged Children with Perinatal HIV Exposure"

**Supplemental Table 1. Comparison of cross-sectional linear regression and longitudinal GEE models for dietary frequency predictors of weight-for-age Z-scores (WAZ).**

| Predictor (vs Rarely) | LM β (95% CI), p | GEE β (95% CI), p |
|---|---|---|
| Eggs (Weekly) | **0.76 (0.05–1.47), *p*= 0.035** | 0.17 (–0.26–0.60), *p*= 0.44 |
| Eggs (Daily) | **0.94 (0.17–1.71), *p*= 0.018** | 0.19 (–0.31–0.68), *p*= 0.46 |
| Beans (Weekly) | –0.55 (–1.36–0.27), *p*= 0.19 | –0.49 (–1.00–0.02), *p*= 0.06 |
| Beans (Daily) | –0.90 (–1.86–0.06), *p*= 0.07 | **–0.74 (–1.36—0.13), *p*= 0.018** |
| Meat (Weekly) | 0.12 (–0.29–0.53), *p*= 0.56 | 0.09 (–0.16–0.34), *p*= 0.46 |
| Vegetables (Weekly) | –0.10 (–0.87–0.68), *p*= 0.80 | –0.03 (–0.66–0.60), *p*= 0.93 |
| Vegetables (Daily) | –0.10 (–0.94–0.75), *p*= 0.82 | 0.00 (–0.69–0.70), *p*= 1.00 |
| Multivitamins (Weekly) | –0.34 (–0.99–0.31), *p*= 0.31 | –0.02 (–0.38–0.33), *p*= 0.91 |
| Multivitamins (Daily) | –0.07 (–0.40–0.26), *p*= 0.68 | 0.00 (–0.27–0.27), *p*= 1.00 |
